# High Frequency of Repeat Expansions in UK Biobank Suggests Protective Rare Variants Modulate C9ORF72 Penetrance in Neurodegeneration

**DOI:** 10.1101/2025.09.18.25336048

**Authors:** E Molinari, A Nickerson, J Quistrebert, A Kless

## Abstract

Short tandem repeat expansions are significant contributors to human disease and several repeat-carrying loci have been identified as responsible for severe neurological disorders. Recent population-based studies showed that the frequency of repeat expansion variants is considerably higher than the prevalence of the disease they cause, suggesting that additional genetic, epigenetic, or environmental factors may influence penetrance and clinical manifestation. This is expected in the case of severe, adult-onset disorders where penetrance is age-dependent, and the incidence is lower than the prevalence. Nonetheless, it remains uncertain whether these variants also exhibit incomplete penetrance in later stages of life. The availability of genetic data linked to longitudinal health records in the UK Biobank allows for direct tracking of disease risk for repeat expansion carriers by age. In our work show that the penetrance of *C9ORF72* repeat expansions, but not that of *HTT* or *CACNA1A*, remains low even late in life and we leverage this feature to identify potential protective variants in ALS.

## Introduction

Short tandem repeats are short, tandemly repeated DNA sequences consisting of 1–6 bp repeated motifs, they constitute 6% of the human genome and have a mutation rate that is orders of magnitude higher than that of the rest of the genome ^1^, contributing strongly to genetic variation. Short tandem repeats are increasingly recognized as important contributors to a wide range of human genetic diseases, with repeat expansions above a certain threshold being considered pathogenic. Alleles with an intermediate repeat length, between normal and pathological, could instead show reduced or variable penetrance and may lead to late-onset or milder phenotypes, or increase disease risk in a context-dependent manner ^2,3^. With the advent of large-scale population-based cohort sequencing efforts, it emerged that several disease-linked repeat expansions in the pathological range are more frequent in the general population than the disease they cause ^4–8^. Several hypotheses can be formulated to explain this unexpected finding. Disorders associated with these mutations may be underdiagnosed or misdiagnosed. Additionally, somatic mosaicism could result in an overrepresentation of long repeat alleles in blood compared to the central nervous system (CNS) and repeat expansion variants may also exhibit incomplete penetrance. Moreover, for certain severe, adult-onset disorders, it is indeed expected that the frequency of carriers of the mutant allele should exceed the reported disease prevalence, whereby, at a certain point in time, some carriers are simply not old enough to have developed the disease, and the prevalence is low compared to incidence, due to short survival.The UK Biobank is a large-scale biomedical database characterised by the availability of longitudinal data, which makes this cohort uniquely suited to directly track the disease risk by age to assess the cumulative risk of being diagnosed with a certain disease. Here we leverage UK Biobank data to complement the observation that the frequency of carriers exceeds the prevalence of the disease, by assessing the incidence of the disease by age, among carriers of repeat expansion variants linked to neurodegeneration. Further, we show that only a fraction of carriers of repeat expansions in the *C9ORF72* locus are affected by the associated diseases even late in life and preliminarily leverage the low penetrance of this mutation to identify potential genetic modifiers.

## Results

### Unexpected high frequency of repeat expansion carriers confirmed in the UK Biobank

For our analysis, we selected a subset of loci that carry short tandem repeats which, when expanded, cause neurodegenerative disorders in humans and for which ExpansionHunter length estimates have been shown to be accurate ^9,10^ (Table 1). Following the release of whole genome sequencing (WGS) data from half a million UK Biobank volunteers, we interrogated the ExpansionHunter output to calculate the number of individuals that carry a repeat length that is above the threshold considered to be pathological for each locus in our list (Table 1 and Figure 1).

**Table 1.** List of loci included in the study that carry short tandem repeats which, when expanded, cause neurodegenerative disorders and prevalence of the associated disease. ^*^Repeat-size thresholds according to Ibanez et al ^8^; ^**^Prevalence among Europeans or the highest prevalence value reported by epidemiological studies, unless due to a known founder effect

| Gene | Type of repeats | Intermediate range | Pathological threshold* | Disease | Disease prevalence per 100,000** |
| --- | --- | --- | --- | --- | --- |
| AR | CAG in exon 1 | 35-37 | >37 | Bulbospinal muscular atrophy (BSMA) | 3.33 <sup>7</sup> |
| ATN1 | CAG in exon 5 | 35-47 | >47 | Dentatorubral pallidoluysian atrophy (DRPLA) | 0.70 <sup>11</sup> |
| ATXN1 | CAG in exon 8 | 39-43 | >43 | Spinocerebellar ataxia type 1 (SCA1) | 2.00<sup>12</sup> |
| ATXN2 | CAG in exon 1 | 33-34 | >34 | Spinocerebellar ataxia type 2 (SCA2) | 0.90<sup>13</sup> |
| ATXN3 | CAG in exon 8 | 45-59 | >59 | Spinocerebellar ataxia type 3 (SCA3) | 2.50<sup>13</sup> |
| ATXN7 | CAG in exon 1 | 34-36 | >36 | Spinocerebellar ataxia type 7 (SCA7) | 0.33<sup>14</sup> |
| C9ORF72 | GGGGCC in intron 1 | NA | >30 | Amyotrophic lateral sclerosis (ALS) | 1.20<sup>15</sup> |
| CACNA1A | CAG in exon 7 | 19 | >19 | Spinocerebellar ataxia type 6 (SCA6) | 0.75<sup>13</sup> |
| HTT | CAG in exon 1 | 36-39 | >39 | Huntington disease (HD) | 9.71<sup>16</sup> |
| TBP | CAG/ CAA in exon 3 | 41-48 | >48 | Spinocerebellar ataxia type 17 (SCA17) | 0.16<sup>17</sup> |

**Figure 1.**
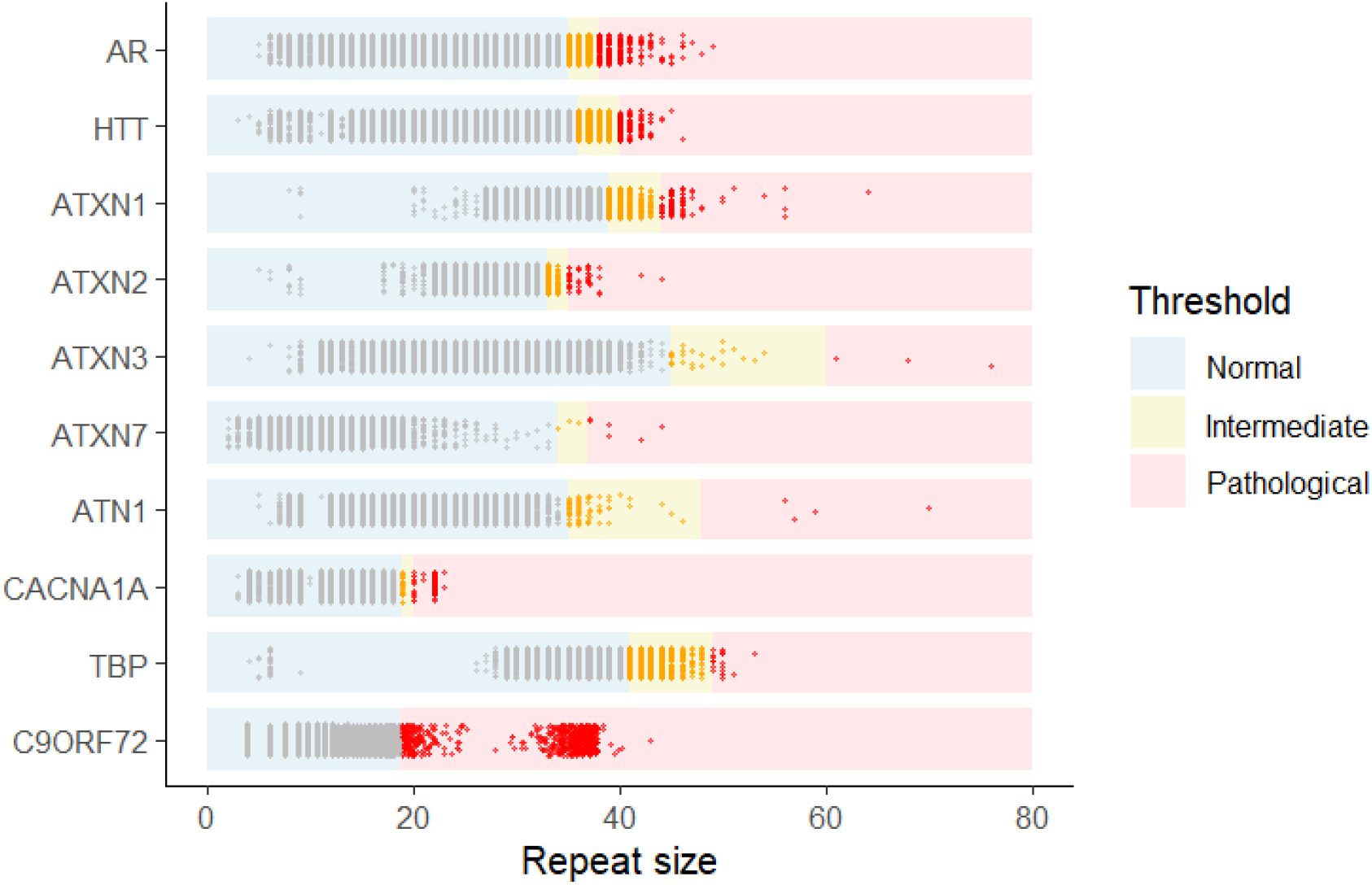
Distribution of repeat size from available genomes in the UK Biobank for each locus considered on this study and as estimated by ExpansionHunter. Each dot corresponds to the long allele for each locus in each genome. Normal, intermediate and pathological repeat size thresholds for each locus, as per Ibanez et al. ^8^, are represented here as shaded background.

We then compared the frequency of repeat expansion carriers in the UK Biobank with the frequencies reported by Ibanez et al ^8^ and by Gardiner et al ^5^, as well as with the prevalence of the disease caused by each of the loci analyzed, as reported by previous epidemiological studies (Table 1, Figure 2 and Supplementary Figure 1a). It should be noted that, for certain loci, there is no consensus around the repeat-size thresholds to be considered intermediate or pathological. For an efficient comparison, we utilized the repeat-size thresholds for pathological repeat expansions reported and used by Ibanez or Gardiner ^5,8^ (Table 1 and Supplementary Figure 1b). Moreover, there is ample geographical variability for the prevalence of some of the disorders associated with the loci considered in this study. Where possible, we use prevalence estimates for Europe or the highest value reported when estimates are extremely variable, unless due to a known founder effect. Overall, there is good agreement between the frequency of repeat expansion carriers in the UK Biobank and the values reported by Ibanez and Gardiner ^5,8^. Strikingly, for the loci analyzed, repeat expansion carriers are up to 115- and 2600-fold (depending on the repeat-size thresholds used) more frequent in the UK Biobank than the reported prevalence of the disease they cause (Figure 2 and Supplementary Figure 1a). We have also looked at the number of carriers of repeat expansion that had a relevant ICD code reported in their health records; that is those that had been diagnosed with the disease caused by the repeat expansion mutation that they carry. For all analyzed loci, only a small proportion of UK Biobank repeat expansion carriers had a relevant ICD code associated with their health records (Table 2).

**Table 2.** Number of carriers of pathological repeat expansions for each locus and number of carriers diagnosed with the associated condition/outcome in the UK Biobank. ^*^Repeat-size thresholds according to Ibanez et al ^8^. For AR, number of males only repeat carriers is reported here.

| Gene | Medical condition (ICD-10 code)/algorithmically defined outcome | Number of carriers of pathological repeat expansion in the UK Biobank* | Number of carriers diagnosed with associated condition/outcome in the UK Biobank | Percentage of carriers of pathological repeat expansion that have been diagnosed with the associated condition/outcome in the UK Biobank |
| --- | --- | --- | --- | --- |
| AR | Motor neuron disease | 87** | 0 | 0% |
| ATN1 | G11 | 4 | 1 | 25% |
| ATXN1 | G11 | 106 | 0 | 0% |
| ATXN2 | G11 | 44 | 4 | 9% |
| ATXN3 | G11 | 3 | 3 | 100% |
| ATXN7 | G11 | 6 | 2 | 33% |
| C9ORF72 | Motor neuron disease | 676 | 51 | 7% |
| C9ORF72 | Frontotemporal dementia | 676 | 16 | 2% |
| CACNA1A | G11 | 77 | 20 | 26% |
| HTT | G10 | 163 | 45 | 28% |
| TBP | G11 | 20 | 0 | 0% |

**Figure 2.**
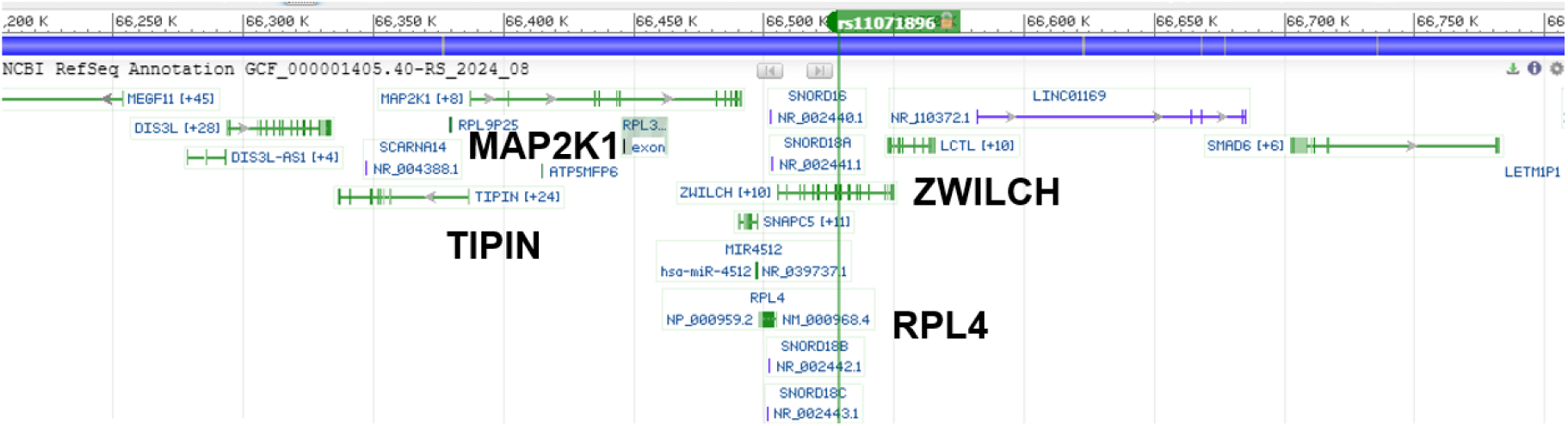
Region of chromosome 15 with clustering of variants enriched in the *C9ORF72*-HRE ALS^−^ group.

**Figure 2.**
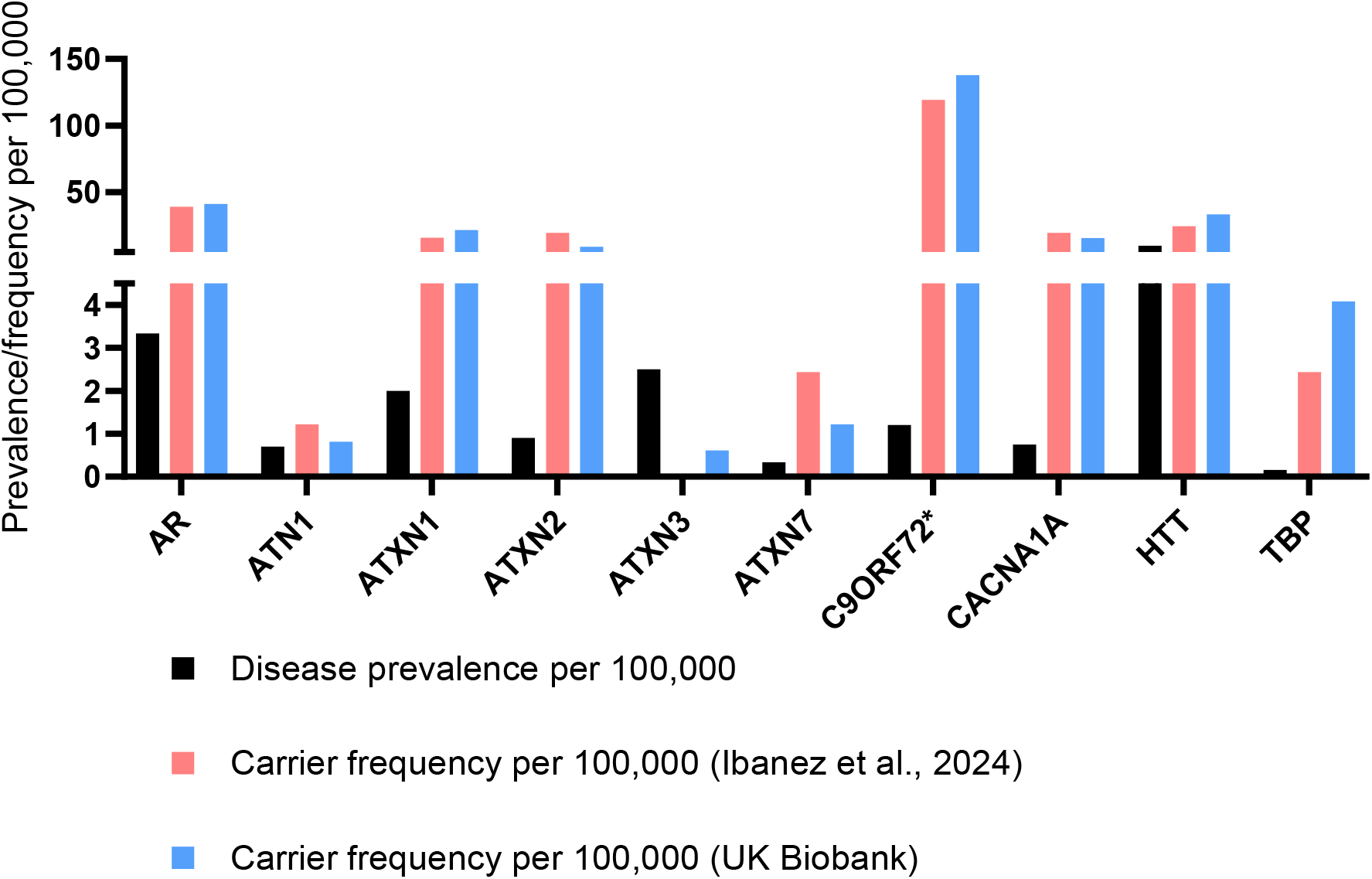
Frequency in the UK Biobank of carriers of pathological repeat expansions (according to repeat-size thresholds reported by Ibanez et al ^8^), in loci associated with neurodegenerative disorders. Frequency of carriers of pathological repeat expansions in the UK Biobank are compared with the reported prevalence of the associated neurodegenerative disorders and with the frequency of carriers of pathological repeat expansions in TOPMED and 100,000 Genomes Project as reported by Ibanez et al ^8^. AR frequency in the UK Biobank refers to male carriers only. ^*^The prevalence reported here refers to ALS only.

### The cumulative risk for repeat expansion carriers of being diagnosed with the associated disease remains low late in life in the case of *C9ORF72*

Some of the diseases examined in this study are adult-onset and highly severe. Consequently, they exhibit an age-dependent increase in risk and a low prevalence that does not reflect their incidence. Therefore, it is expected that the frequency of individuals carrying the mutant allele surpasses the reported prevalence of the disease. For these reasons, we aimed to estimate the age-dependent cumulative risk of disease diagnosis among repeat expansion carriers, providing a more accurate measure of the penetrance associated with these mutations. It should be noted that penetrance estimates may be skewed downwardly by the “healthy volunteer” participation bias that characterizes the UK Biobank, as other volunteer-based cohorts, whereby participants tend to be healthier than the population from which they were sampled ^18^. Nevertheless, it has been shown that, in other volunteer-based cohorts, this bias attenuates over time, due to the development of chronic diseases as the cohort ages ^19–21^. Similarly, we reasoned that the participation bias will be milder when considering health outcomes that are typically associated with an age of onset greater than the minimum age cutoff for recruitment in the cohort.

Thus, to mitigate a potential “healthy volunteer effect”, we focused on a smaller subset of loci that cause disorders with an average age of onset greater than 40 years, i.e. the minimum age cutoff for recruitment in the UK Biobank (*C9ORF72, CACNA1A* and *HTT*). The mean age of onset of the motor neuron disease bulbospinal muscular atrophy (BSMA), caused by a CAG repeat expansion in exon 1 of the *AR* gene, is also above 40 (43 years) ^7^.Remarkably, out of the 87 carriers of pathological repeat expansions in AR, none was associated with a motor neuron disease in the UK Biobank. Consequently, we could not plot the penetrance of the expansion mutation in function of age for this locus. Our analysis shows that, while with different trends, the cumulative risk for repeat expansion carriers increases with age as expected. For both the *CACNA1A* and the *HTT* loci, the cumulative risk for repeat expansion carriers of being diagnosed with the associated disease is around 70% by 82 years (Figure 3b,c). Strikingly, repeat expansion mutations in the *C9ORF72* locus may be incompletely penetrant even late in life, with a cumulative risk of being diagnosed with ALS and/or FTD of only 25% at 80 years or older (Figure 3a). This is consistent with the point-penetrance estimates from previous studies that used for their calculations complementary methods based on population-scale data ^22,23^. The length of the repeat expansion has been reported, in some cases, to be a strong modifier of the age of onset ^24,25^. Consistent with what has been reported by several studies ^24,26^, repeat length in the case of *HTT* appears to be inversely correlated with the age of onset in the UK Biobank dataset, while in the case of *C9ORF72* the relationship between length and age of onset remains debated ^27^ and is not evident in our dataset (Supplementary Figure 3a,b,d). The repeat length in *CACNA1A* is known to show a strong inverse correlation with age of onset ^28^, which is not evident in our dataset, probably due to the fact that the affected carriers in the UK Biobank have repeats that cluster in the extremely tight size range of 22-23, with most of them measuring 22 (Supplementary Figure 3c,d). We also tested whether short repeat expansions were enriched among carriers that remained symptom-free and show that indeed repeat length appears to be a modifier of penetrance (Supplementary Figure 4).

**Figure 3.**
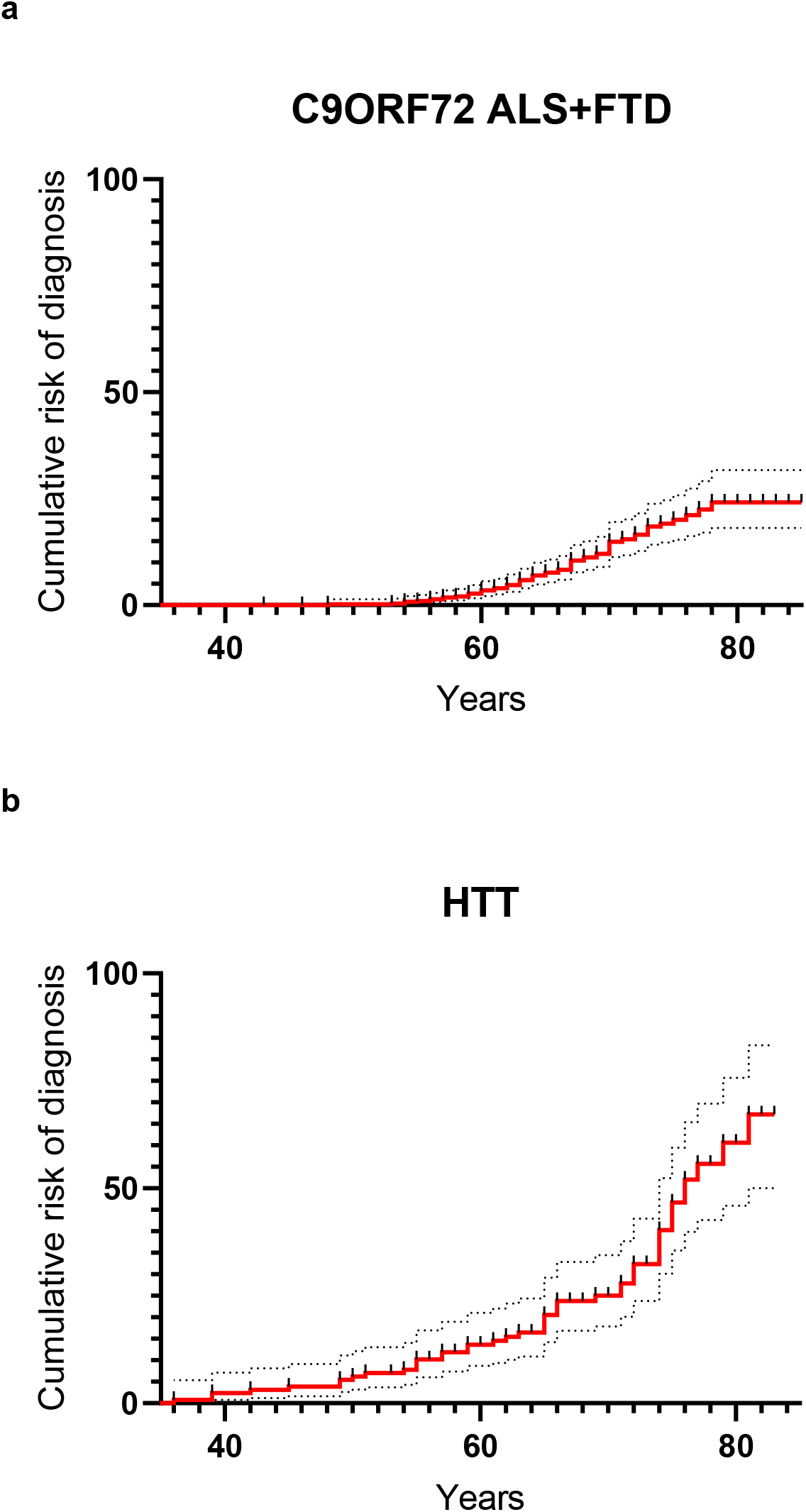
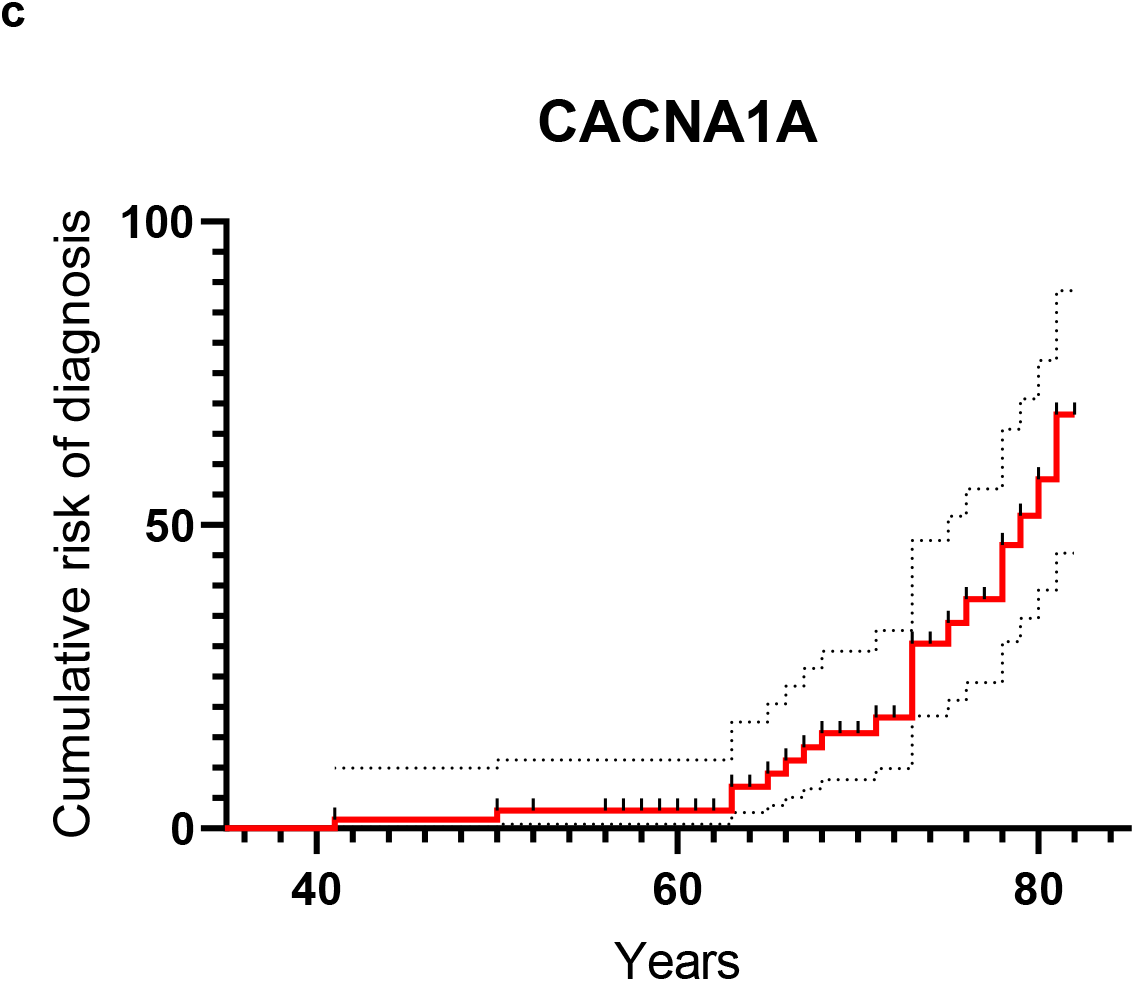
Cumulative risk for repeat expansion carriers of being diagnosed with the associated disease. **(a)** Cumulative risk for carriers of repeat expansions in the *C9ORF72* locus to develop ALS and/or FTD; **(b)** cumulative risk for carriers of repeat expansions in the *HTT* locus to develop HD; **(c)** cumulative risk for carriers of repeat expansions in the *CACNA1A* locus to develop SCA.

### Leveraging the reduced penetrance of *C9ORF72* repeat expansion to identify genetic modifiers of *C9ORF72-*ALS disease risk

Reduced penetrance of the *C9ORF72* repeat expansion mutation may suggest the presence of additional genetic or environmental factors that contribute to disease risk. In fact, additional contributors have been shown to act on top of monogenic variants to modify the disease risk ^29,30^ and, while the presence of the *C9ORF72* repeat expansion reduces the number of steps required for disease development, other factors also contribute to the clinical presentation of the disease ^31^. In order to identify potential genetic contributors to ALS that act on top of the *C9ORF72* repeat expansion, we carried out a variant enrichment analysis by comparing the frequency of ~27,000,000 moderate- and high-impact variants from the whole exome sequencing (WES) available in the UK Biobank of *C9ORF72* repeat expansion carriers that were diagnosed with ALS (*C9ORF72*-HRE ALS+) and the carriers that remained symptom-free (*C9ORF72*-HRE ALS-). We identified a locus in the region q22.31 of chromosome 15 where several variants enriched in the *C9ORF72*-HRE ALS^−^ group appeared to cluster (Table 3, Supplementary Table 1). This region spans the genes *ZWILCH, TIPIN, MAP2K1* and *RPL4* (Figure 4).

**Table 3.**
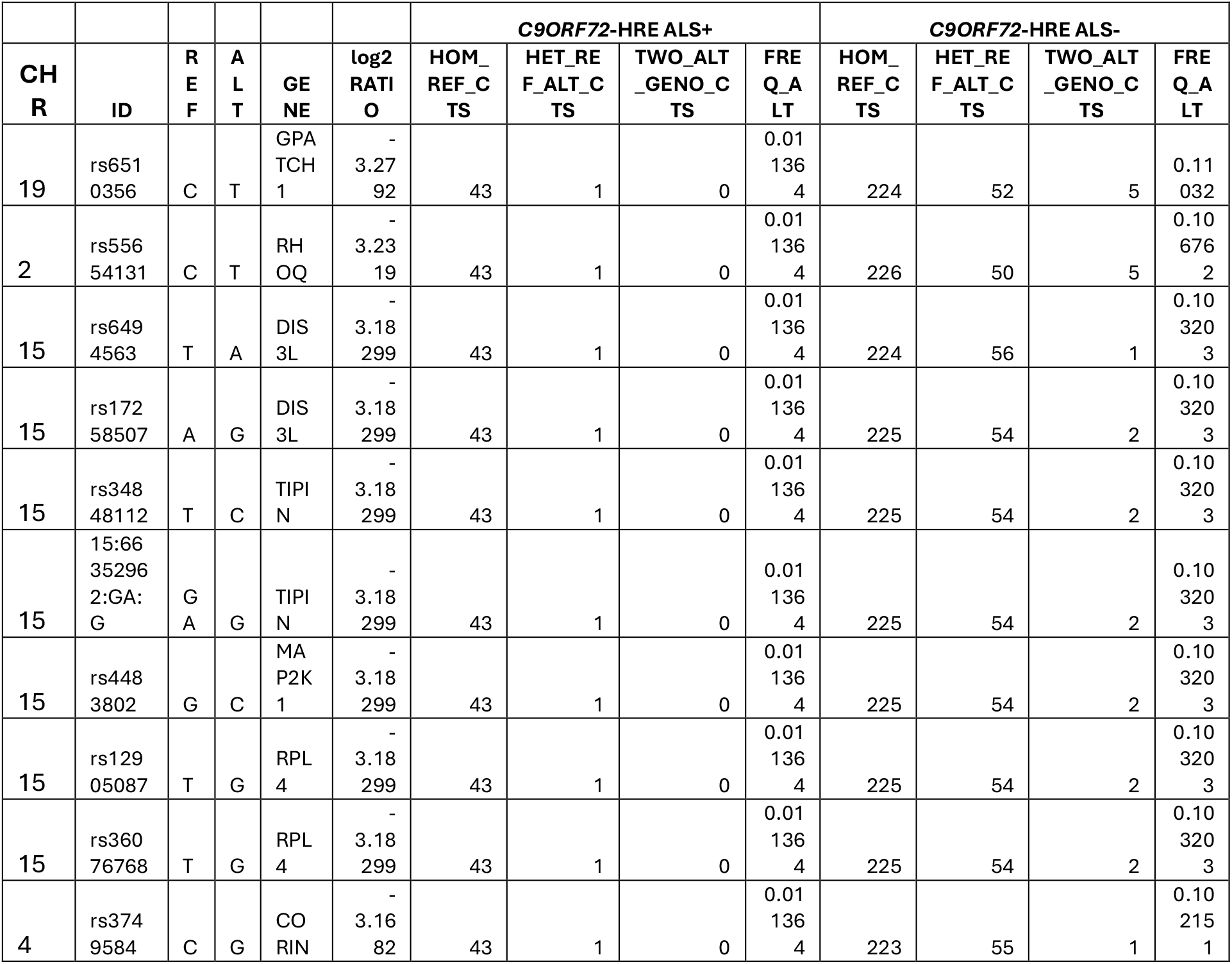
Top 10 enriched variants in the *C9ORF72*-HRE ALS^−^ group vs the *C9ORF72*-HRE ALS^+^ group. CHR, Chromosome; ID, Variant ID; REF, Reference allele; ALT, Alternative allele; GENE, Gene symbol; log2 RATIO, log2 of the ratio between the alternative allele frequency in the *C9ORF72*-HRE ALS^+^ group and the alternative allele frequency in the *C9ORF72*-HRE ALS^−^ group; HOM_REF_CTS, Reference allele homozygous counts; HET_REF_ALT_CTS, Heterozygous counts; TWO_ALT_GENO_CTS; Alternative allele homozygous counts; FREQ_ALT, Frequency of the alternative allele. (Autosomal variants only).

While our analysis is generally underpowered due to the low number of individuals, we tried to substantiate these findings by cross-referencing them with a genetic association analysis using the WES from 665 motor neuron disease individuals (ICD-10 12.2) and two controls cohorts with 158K and 15K participants in the UK Biobank to identify putative protective rare variants. We found several missense variants that showed an odd ratio that is far below 1 which tells that the minor allele is more frequent in the control group and therefore protective. Such protective missense variants can also be experimentally validated by site directed mutagenesis. Since the discovery of protective rare variants depends on the control group we have performed these genetic associations in two groups of different size to account for the variability. Most of our genetic associations were not present in both analysis but intriguingly, one variant in the same region of chromosome 15 (rs11071896), mapping on the ZWILCH gene was present in both genetic associations (Table 4a, Figure 4). Additionally, we have found a protective 3’-UTR variant in C9orf72 rs3849944 which regulates gene expression of C9orf72 in brain tissue. We further investigated the C9orf72 missense variant rs17769294 (Asn207Ser) in a cohort of 71 individuals carrying pathogenic repeat expansions (>100 repeats) associated with motor neuron disease (Table 4b). Leveraging a genome-wide epistatic interaction analysis across 1.5 million missense variants from the UK Biobank, we identified several statistically significant interactions that may act as genetic modifiers influencing disease susceptibility and progression. Among these, a particularly strong protective interaction was observed with the zinc-finger protein ZNF577 (rs150301895, Ser71Leu), previously implicated in DNA methylation and mRNA expression regulation (Lorenzo PM et al., Front. Endocrinol. 11:245). This interaction yielded a chi-square statistic of 17.862, an odds ratio (OR) of 0.199, and a beta coefficient of −1.612, indicating a substantial reduction in disease risk for carriers of both variants. Conversely, several interactions were associated with increased disease risk. A missense variant in CROCC (rs147926226, Arg1908Cys) exhibited a high-risk interaction (OR = 10.284, beta = 2.331), suggesting a synergistic disruption of cytoskeletal integrity and intracellular trafficking which are processes critical for neuronal maintenance. Similarly, C3orf20 (rs148474695, Gly112Glu) showed an even stronger interaction (OR = 11.348, beta = 2.429), potentially influencing transcriptional or epigenetic mechanisms that intersect with C9orf72-related pathways such as RNA metabolism and stress granule dynamics. Additionally, UBE4B (rs61760196, Leu442Phe) emerged as a significant interactor (OR = 2.961, beta = 1.085), possibly contributing to disease pathology through its role in ubiquitination and proteostasis regulation. All identified interactions remained significant after multiple testing correction (Benjamini-Hochberg adjusted p-values < 0.05), underscoring their robustness. The observed spectrum of effect sizes from highly protective (OR < 0.2) to strongly deleterious (OR > 10) highlights the complex genetic architecture of C9orf72-associated motor neuron disease and points toward novel avenues for therapeutic intervention targeting protective modifiers.

**Table 4a:**

| CHR | POS | ID | REF | ALT | Controls | OR | SE | Z_STAT | P | P_BH | Gene | AA-exchange |
| --- | --- | --- | --- | --- | --- | --- | --- | --- | --- | --- | --- | --- |
| 15 | 66528912 | rs11071896 | A | G | 15096 | 0.635 | 0.105 | -4.32 | 1.56E-05 | 7.70E-05 | ZWILCH | p.Ser344Gly |
| 15 | 66528912 | rs11071896 | A | G | 158141 | 0.735 | 0.079 | -3.899 | 9.64E-05 | 3.10E-04 | ZWILCH | p.Ser344Gly |
| 9 | 27560596 | rs3849944 | T | C | 172056 | 0.373 | 0.209 | -4.716 | 2.40E-06 | 4.24E-06 | C9orf72 | 3_prime_UTR |

**Table 4b:**

| Gene A | ID_A | AA-exchange A | Gene B | ID_B | AA-exchange B | CHISQ | OR | P | P_BH | beta |
| --- | --- | --- | --- | --- | --- | --- | --- | --- | --- | --- |
| C9orf72 | rs17769294 | p.Asn207Ser | ZNF577 | rs150301895 | p.Ser71Leu | 17.862 | 0.199 | 2.38E-05 | 3.18E-03 | -1.612 |
| C9orf72 | rs17769294 | p.Asn207Ser | CROCC | rs147926226 | p.Arg1908Cys | 16.706 | 10.284 | 4.37E-05 | 3.18E-03 | 2.331 |
| C9orf72 | rs17769294 | p.Asn207Ser | UBE4B | rs61760196 | p.Leu442Phe | 15.619 | 2.961 | 7.75E-05 | 3.72E-03 | 1.085 |
| C9orf72 | rs17769294 | p.Asn207Ser | C3orf20 | rs148474695 | p.Gly112Glu | 15.576 | 11.348 | 7.93E-05 | 3.72E-03 | 2.429 |

## Discussion

Here we show that the frequency of pathological repeat expansion variants associated with neurodegenerative disorders is high in the UK Biobank; up to 115-fold higher than the prevalence of the associated disease. The frequencies we observed in the UK Biobank are largely in agreement with the frequencies reported by other recent studies that used population-based cohorts, mainly of European descent ^4–8^. It is commonly accepted that a deleterious genotype should be no more prevalent in the population than the disease it causes, but this position is becoming increasingly challenged with several low penetrance variants being identified in large-scale studies ^32–35^. Moreover, in the case of severe, adult-onset disorders, where the risk of developing the disease is age-dependent and the survival short, the frequency of the deleterious genotype is indeed expected to be higher than the prevalence of the associated disease. Whether these variants are only apparently incompletely penetrant, with potential full penetrance late in life, it is unclear. The availability in the UK Biobank of genetic data linked to longitudinal health records allowed us to directly track the risk of disease by age for repeat expansion carriers in the *C9ORF72, HTT* and *CACNA1A* loci. By doing so, we were able to show that, as expected, the risk of disease increases by age, but it follows different trends for each locus. Moreover, while in the case of *HTT* and *CACNA1A* the penetrance is high late in life, reaching 70% by 82 years, the risk of ALS and FTD combined for *C9ORF72* repeat expansion carriers remains low (25%) even late in life. Low penetrance of the *C9ORF72* repeat expansion would be in agreement with the incomplete disease penetrance observed in pedigrees associated with this expansion ^36^, with the apparently sporadic presentation of *C9ORF72*-ALS/FTD cases and with recent point-penetrance estimates for *C9ORF72* expansions calculated based on population-scale data ^22,23^. However, our penetrance estimates are also in stark disagreement with previous attempts to calculate the age-dependent penetrance of *C9ORF72* repeat expansion, which reported near fully penetrant estimates by 80 years of age ^37^. However, previous analyses likely yield biased estimates due to their reliance on disease cohorts, which are inherently subject to high ascertainment bias. Remarkably, with a limited ascertainment bias and the availability of longitudinal data, the UK Biobank appears to be a unique fit for cumulative risk analyses, and is also becoming an increasingly elderly cohort, therefore particularly suited for the study of late onset neurodegenerative disorders. While uniquely suited for this type of analysis, the use of the UK Biobank made in this study is not without limitations. The UK Biobank is a relatively uniform cohort of European descent, meaning that the prevalence estimates found in this study cannot be extended to individuals of a different descent. Moreover, the UK Biobank cannot be considered fully representative of the general population, as there appears to be a “healthy volunteer” bias in the recruitment of participants ^18^, that could lead to an underestimation of penetrance. However, the impact of such bias likely remains limited when considering adult-onset, short-survival disorders like ALS and HD. While the likelihood of an individual being recruited to the UK Biobank while being affected by such diseases may be low, the prevalence at any time point is sadly extremely low due to the short survival associated with these diseases, limiting the impact of this bias on our penetrance estimates. Another limitation of this study is that we relied on short-read sequencing to estimate the length of the repeats. While long-read sequencing would be a more accurate, direct method for this analysis, it is currently not available for UK Biobank genomes and the accuracy of the ExpansionHunter algorithm has been demonstrated for the loci analyzed in this study ^9,10^. Finally, it should be acknowledged that alternative or complementary hypotheses other than low mutation penetrance can be formulated to explain the fact that only a small proportion of *C9ORF72* repeat expansion carriers are diagnosed with the disease. For instance, due to the somatic instability of tandem repeats, which is generally considered to be a driver in the progression and severity of some repeat expansion disorders ^38^, we cannot exclude that certain carriers of repeat expansions in blood may have repeat sizes below the pathological threshold in the CNS. Moreover, it is possible that some of the carriers are indeed affected by the disease but have not been diagnosed as such. However, for the disorders that are object of this study, while late diagnosis is common, underdiagnosis is unlikely, considering the severity of these disorders. To address the possibility that some of the undiagnosed carriers have in fact simply been misdiagnosed, we actively filtered out any adjacent diagnosis in the generation of our non-diagnosed cohorts (see Methods). Incomplete penetrance of the repeat expansion variants, on the other hand, appears to be a valid explanation, especially in light of the increased recognition that many monogenic disease-implicated variants have lower penetrance in the general population than was originally estimated using disease cohorts ^32–35^. The potential incomplete penetrance of repeat expansion variants could have several important implications. First, incomplete penetrance of repeat expansion variants could have an important impact on the genetic counselling practices for repeat expansion carriers and their families. Second, incomplete penetrance could make clinical trials on pre-symptomatic mutation carriers intractable. Finally, incomplete penetrance could be leveraged to identify potential modifiers that protect healthy carriers. Here, we provide a preliminary attempt to leverage the low penetrance of the *C9ORF72* repeat expansion mutation to identify additional genetic factors that modify the disease risk. The number of *C9ORF72* repeat expansion carriers in the UK Biobank, although unexpectedly high, does not allow a sufficiently powered genome-wide association study (GWAS) for the identification of these modifiers, so we limited our preliminary analysis to a simple comparison of the frequency of moderate and high-impact autosomal coding variants between the *C9ORF72*-HRE ALS^−^ group and the *C9ORF72*-HRE ALS^−^ group. To substantiate the results of our variant enrichment analysis, we cross-referenced them with the results of an EWAS that we carried out on UK Biobank ALS cases versus controls to identify potential protective variants. Intriguingly, these two independent analyses converged on a locus on chromosome 15, where several variants, overrepresented in the *C9ORF72*-HRE ALS^−^ group, appeared to cluster and the variant rs11071896 (15q22.31) was also among the most significant hits of the EWAS. This locus spans several genes, including *DIS3L, TIPIN, MAP2K1, RPL4* and *ZWILCH*. Intriguingly, *TIPIN* is a crucial player in DNA damage response ^39^, which is an emerging pathophysiological mechanism of ALS ^40,41^. At this moment, our analysis is not sufficiently powered to confirm the contribution of any of these genes to ALS risk. As public and private investments in sequencing efforts of large cohorts are on the rise, it is conceivable that, in the near future, there will be availability of sequences from enough repeat expansion carriers to run a small GWAS that will allow the identification of genetic modifiers of *C9ORF72*-ALS and other types of ALS, as also single nucleotide variants implicated in ALS have been shown to be incompletely penetrant ^23^. The identification of such genetic modifiers could have a huge impact in the development of novel therapeutics for diseases associated with repeat expansion mutations, such as ALS. ALS is a devastating disease for which sadly no disease-modifying treatment is available, apart from the encouraging case of Tofersen and Jacifusen, two investigational antisense oligonucleotides that showed some promises in the treatment of *SOD1* and *FUS*-associated ALS, respectively ^42–44^. Therapies that tackle the causative gene, like Tofersen, hold a tremendous potential in the treatment of ALS. Nevertheless, our present knowledge of ALS genetics accounts for only a small fraction of sporadic cases, which make up more than 90% of all ALS patients. The identification of protective variants that impact the risk of sporadic ALS could provide an important avenue to develop novel therapies to prevent, halt or slow down the progression of the disease. In conclusion, we propose a shift in the perception of repeat expansions in the *C9ORF72* locus and their role in neurodegeneration. We suggest that repeat expansions in this locus should be regarded as low-penetrance variants; or even as risk alleles, considering the discordant segregation reported in some of the affected families ^35,36^. Our findings have an impact on genetic counselling as well as our understanding of the biology of the associated diseases, hinting at a situation where other factors may play a role, and offering a unique opportunity to identify new players in neurodegeneration.

## Methods

### Genetic association analysis, epistasis, repeat expansion and Annotation

The UK Biobank cohort contains 502,394 participants (54.4% female), aged 37-73 at enrolment (2006-2010).^47^ The age range at the time of this study is 52-89. Participants have undergone genetic sequencing, detailed phenotypic screening, and had their NHS hospital inpatient and prescription data linked. In 2022, the final whole exome sequencing dataset for UKB was released.^48^ We used the UK Biobank Apollo database version for our analysis. The UK Biobank Research Analysis Platform (RAP) was used to perform the analysis of whole exome sequencing data. A generalised linear model regression using the allelic model was performed on the genotype array and whole exome sequencing data using Plink2^46^ using the first 4 principal components, age and sex as covariates. Each of the cohorts were compared to control groups that excludes any diagnosis in neurodegeneration. To correct for the population structure within the UK Biobank we included principal components as covariates in our statistical models. The principal component vectors were calculated using the full PCA mode of Plink2. Further, we excluded the subset of variants in known inversions and MHC regions (chr6:25500000-33500000, chr8:8135000-12000000, and chr17:40900000-45000000) with a minor allele frequency of less than 1% and those within high linkage disequilibrium using the independent pairwise pruning, a 50kb window with steps of 5 as well as an r^2^ threshold of 0.2. Nominally significant genetic variants (−log_10_(p)>5) have been annotated with SNPeff^47^ to identify the gene regions and to understand their impact. To generate testable hypotheses this research emphasizes analysis of rare variants, particularly those involving missense and loss-of-function (LoF) mutations. We examined the epistatic interactions between a particular rare variant and a set of 1.5 million missense and LoF variants across the whole genome. This approach aims to better understand the penetrance of rare variants by genetic modifiers. Additionally, it helps to elucidate the overall genetic architecture that may not be evident when analysing single genetic variants alone. Consideration of epistatic interactions in the results from rare variant genetic associations can improve the accuracy and predictive power of genetic models for complex traits. For risk associations the epistasis was also run to identify potential protective variants. The calculations were performed using SNPint^48^ using the logistic regression approach. For calculating the repeat expansion genotype, we used results from ExpansionHunter^45^ as provided by the UK Biobank RAP platform. We classified repeat expansion carriers as an individual that has a repeat size (REPCN value for the long allele) greater than the pathological threshold reported in Table 1 for each of the loci considered in the study. The confidence in detecting C9ORF72 repeat expansions using short-read sequencing as currently provided by the UK Biobank particularly by the ExpansionHunter algorithm which uses a sequence-graph approach to genotype short tandem repeats (STRs) from PCR-free whole-genome sequencing data is considered moderate to high. ExpansionHunter has been validated against gold-standard methods like RP-PCR and Southern blotting and performs reliably in identifying expansions above the relevant 30-repeat threshold.^45^

### Study population and cohort definition

UK Biobank is a large-scale biomedical database and research resource containing anonymised genetic, lifestyle and health information from UK participants. UK Biobank recruited 500,000 people aged between 40-69 years in 2006-2010. A repeat expansion carrier for a certain locus was considered to have been diagnosed with the associated disease (according to Table 2) if the First occurrences of Health-related outcomes (Category 1712) had a Date of a relevant ICD-10 being first reported not null, or, when an Algorithmically defined outcome was available (motor neuron disease or frontotemporal dementia), the Date of report (p42028 or p42024, respectively) was not null. Since it’s possible that some carriers may have presented with relevant symptoms but could have been misdiagnosed, to generate our non-diagnosed cohorts, we actively filtered out carriers in which the Summary Diagnoses (Data-Field 41270) included ICD-10 codes that could represent any adjacent diagnosis to the one associated with the specific locus according to Table 2. For the *C9ORF72* and the *HTT* loci, we excluded from the non-diagnosed cohort all the carriers that in the Summary Diagnoses field had the following ICD-10 codes: F00, F01, F02, F03, F05, F06.7, G10-G14, G20-26, G30-32, G37, R41.0, R41.3, R41.8. For the *CACNA1A* locus, we excluded from the non-diagnosed cohort all the carriers that in the Summary Diagnoses field had the following ICD-10 codes: F00, F01, F02, F03, F05, F06.7, G10-G14, G20-26, G30-32, G37, G40, R41.0, R41.3, R41.8. We also excluded all the carriers with a null Summary Diagnoses field.

### Statistical analysis

To assess whether there was a correlation between the repeat size and the age of onset, we plotted the age of the carriers that had been diagnosed with the relevant disease, at the Date of the relevant ICD-10 being first reported, by function of the repeat size. We then tried to fit a linear model, using R function lm() and interrogated the coefficients. To assess whether there is a significant difference in the repeat size in function of the disease status, we employed a non-parametric statistical test (Wilcoxon rank sum test), using the R function wilcox.test. The p-values obtained from the genetic association analysis were adjusted using the Benjamini-Hochberg (BH) procedure to control the false discovery rate (FDR). We define results as significant if their BH-adjusted p-value is ≤ 0.05, ensuring that the expected proportion of false discoveries among these findings does not exceed 5%.

### Cumulative disease risk analysis

To generate the Kaplan-Meier curves for graphical representation of the cumulative disease risk for repeat expansion carriers, we filled a Survival table in GraphPad Prism by using the age of the carriers that had been diagnosed with the relevant disease at the Date of the relevant ICD-10 being first reported. We censored outcomes for the carriers that remained symptom-free, based on the age at Date of death (Data Field 40000), or the age in 2022, as Hospital inpatient data, which constitute an important data source for the Algorithmically defined outcomes and the First occurrences categories, had 2022 censoring dates (defined as the last day of the month for which the number of records is greater than 90% of the mean of the number of records for the previous three months). We acknowledge that the date reported in the First occurrences category and the Date of report for the Algorithmically defined outcomes category may not represent the true date of onset of symptoms.

### Variant enrichment analysis

We generated the count files using Plink2 for the two cohorts, *C9ORF72*-HRE ALS^+^ and *C9ORF72*-HRE ALS^−^ and discarded all the variants that had missing counts in >= 10% of the genomes in each cohort. For this preliminary analysis we focused on autosomal chromosomes only. We calculated the frequency of the alternative allele in each allele and generated a log2 RATIO as follows: log2 (FREQ_ALT in the *C9ORF72*-HRE ALS^+^ group/ FREQ_ALT in the *C9ORF72*-HRE ALS^−^ group) and ranked the variants for the highest and lowest log2 RATIO to identify the 10 top enriched variants in the group *C9ORF72*-HRE ALS^+^ group and in the *C9ORF72*-HRE ALS^−^ group, respectively.

## Data Availability

This research has been conducted using data from UK Biobank, a major biomedical database under project ID 85173.

## Acknowledgements

This research has been conducted using data from UK Biobank, a major biomedical database under project ID 85173. The UK Biobank database received ethical approval from the North-West Haydock Research Ethics Committee (REC reference 21/NW/0157) and participants gave informed consent. We would like to thank all participants of the UK Biobank cohort who have provided necessary genetic and phenotypic information.

## Competing interests

EM, AN, JQ and AK are current or former employees of Eli Lilly and Company and may own stock in this company.

## Supplementary figures

**Supplementary Figure 1.**
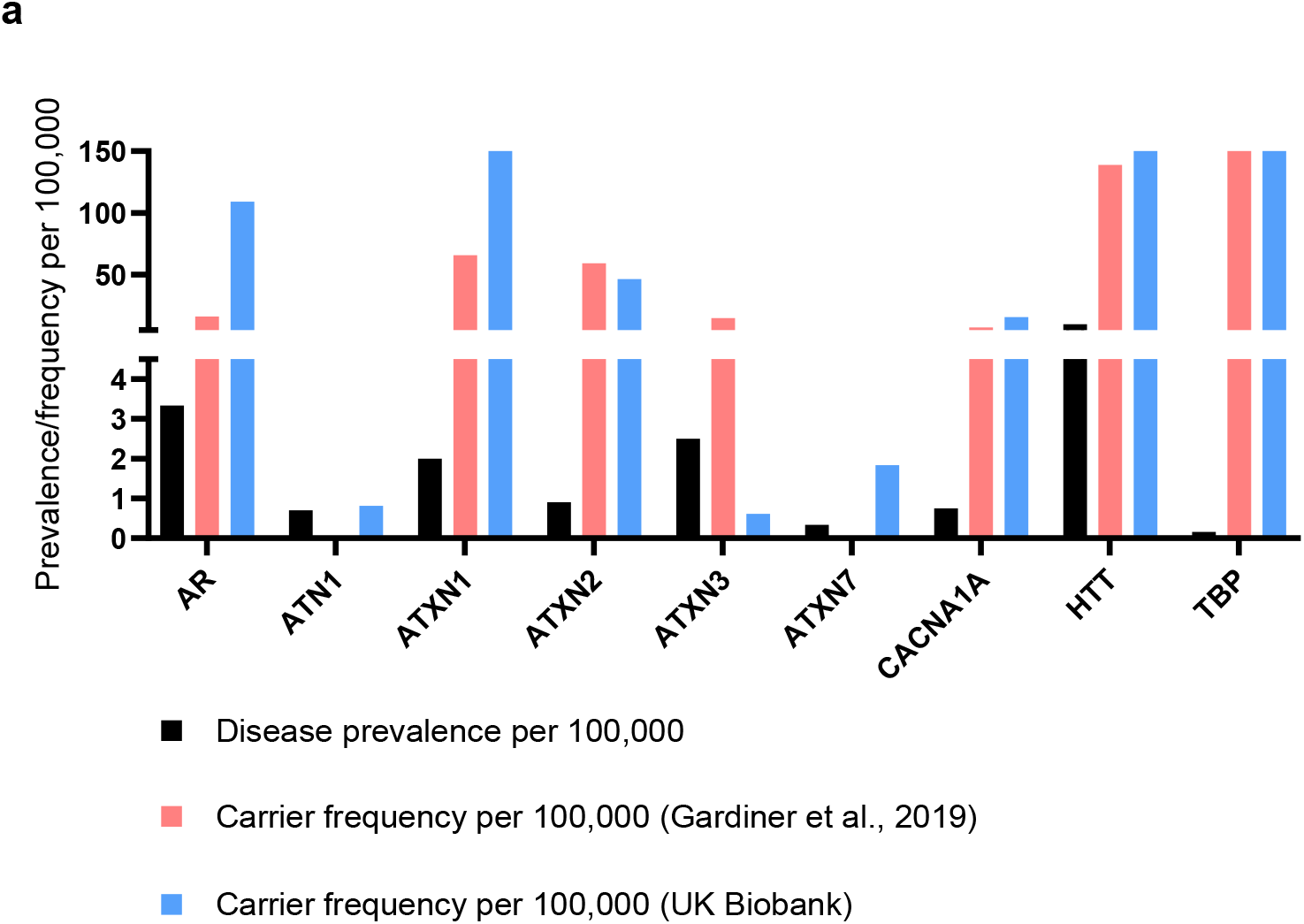
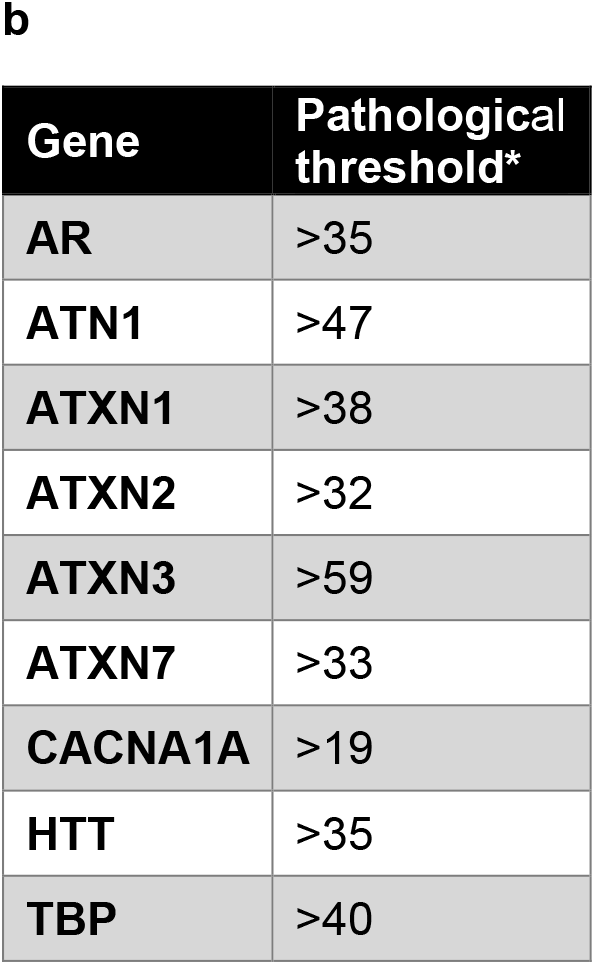
Frequency in the UK Biobank of carriers of pathological repeat expansions (according to repeat-size thresholds reported by Gardiner et al ^5^), in loci associated with neurodegenerative disorders. **(a)** Frequency of carriers of pathological repeat expansions in the UK Biobank are compared with the reported prevalence of the associated neurodegenerative disorders and with the frequency of carriers of pathological repeat expansions in 5 large European population– based cohorts (NESDA, NESDO, NEO, PROSPER, Leiden 85-plus) as reported by Gardiner et al ^5^. AR frequency in the UK Biobank refers to male carriers only. ^*^The prevalence reported here refers to ALS only. **(b)** Pathological thresholds according to Gardiner et al ^5^.

**Supplementary Figure 2.**
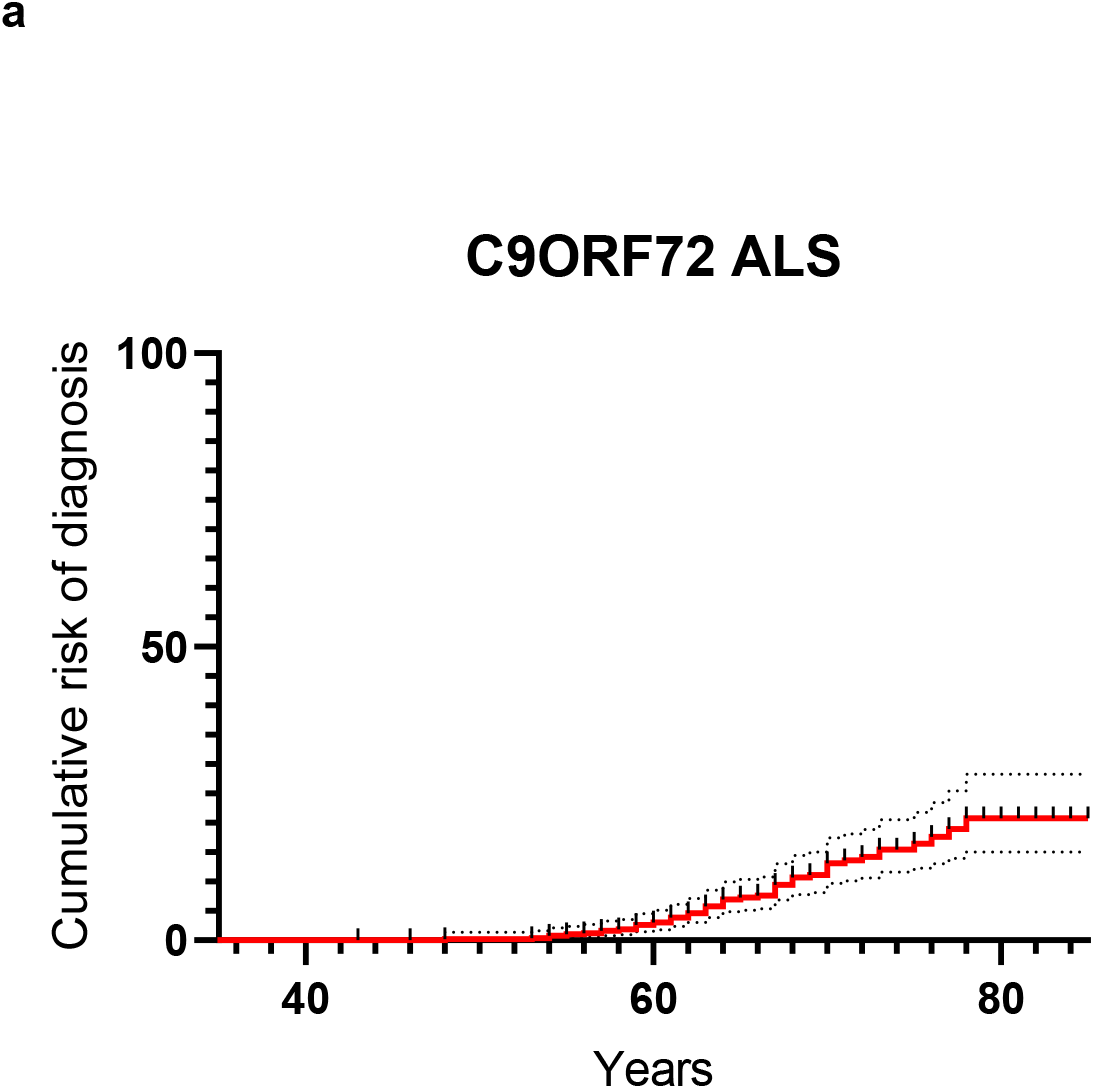
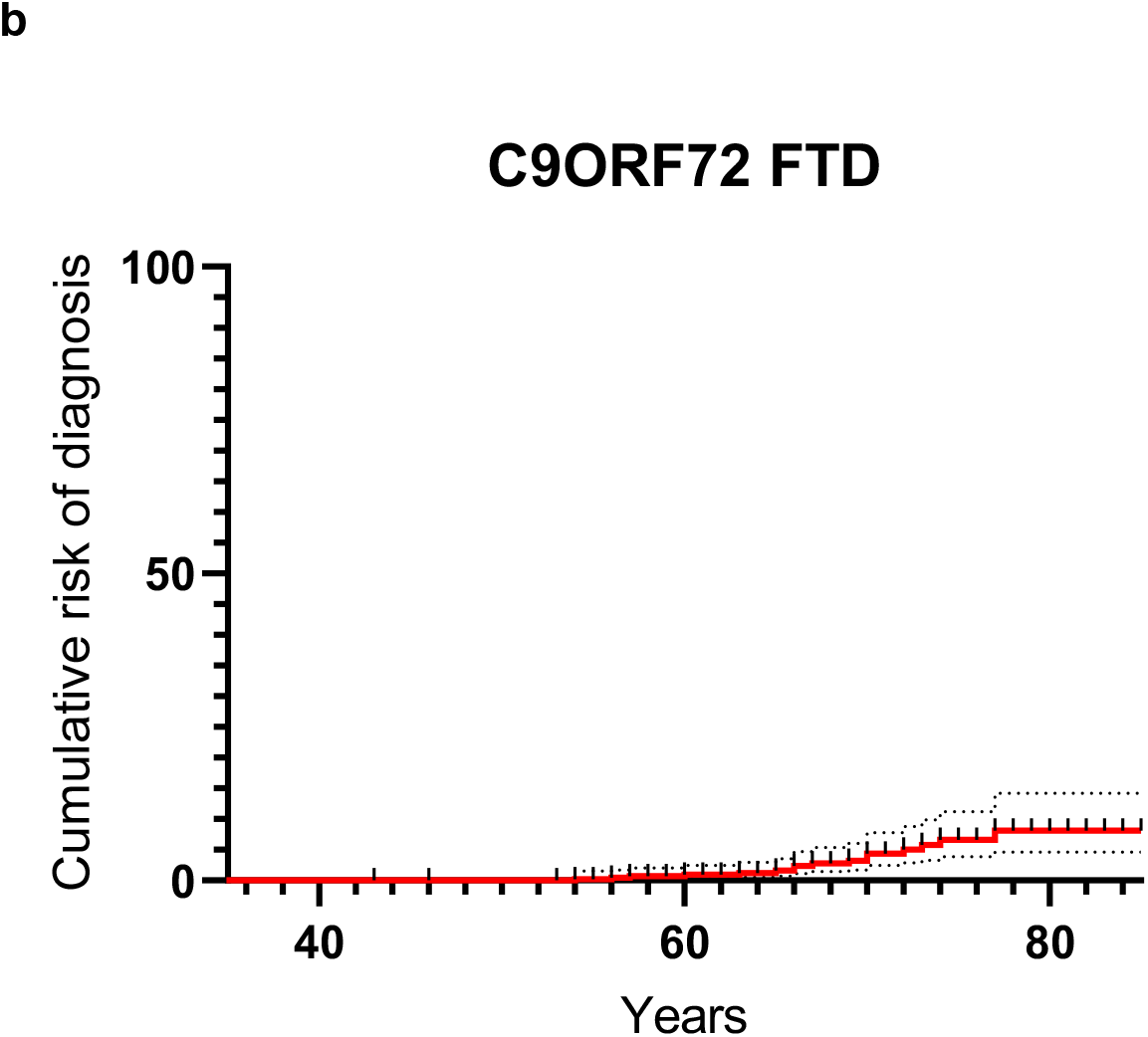
Cumulative risk for ALS and FTD separately in carriers of repeat expansions in the *C9ORF72* locus. **(a)** Cumulative risk for carriers of repeat expansions in the *C9ORF72* locus to develop ALS only; **(b)** cumulative risk for carriers of repeat expansions in the *C9ORF72* locus to develop FTD only.

**Supplementary Figure 3.**
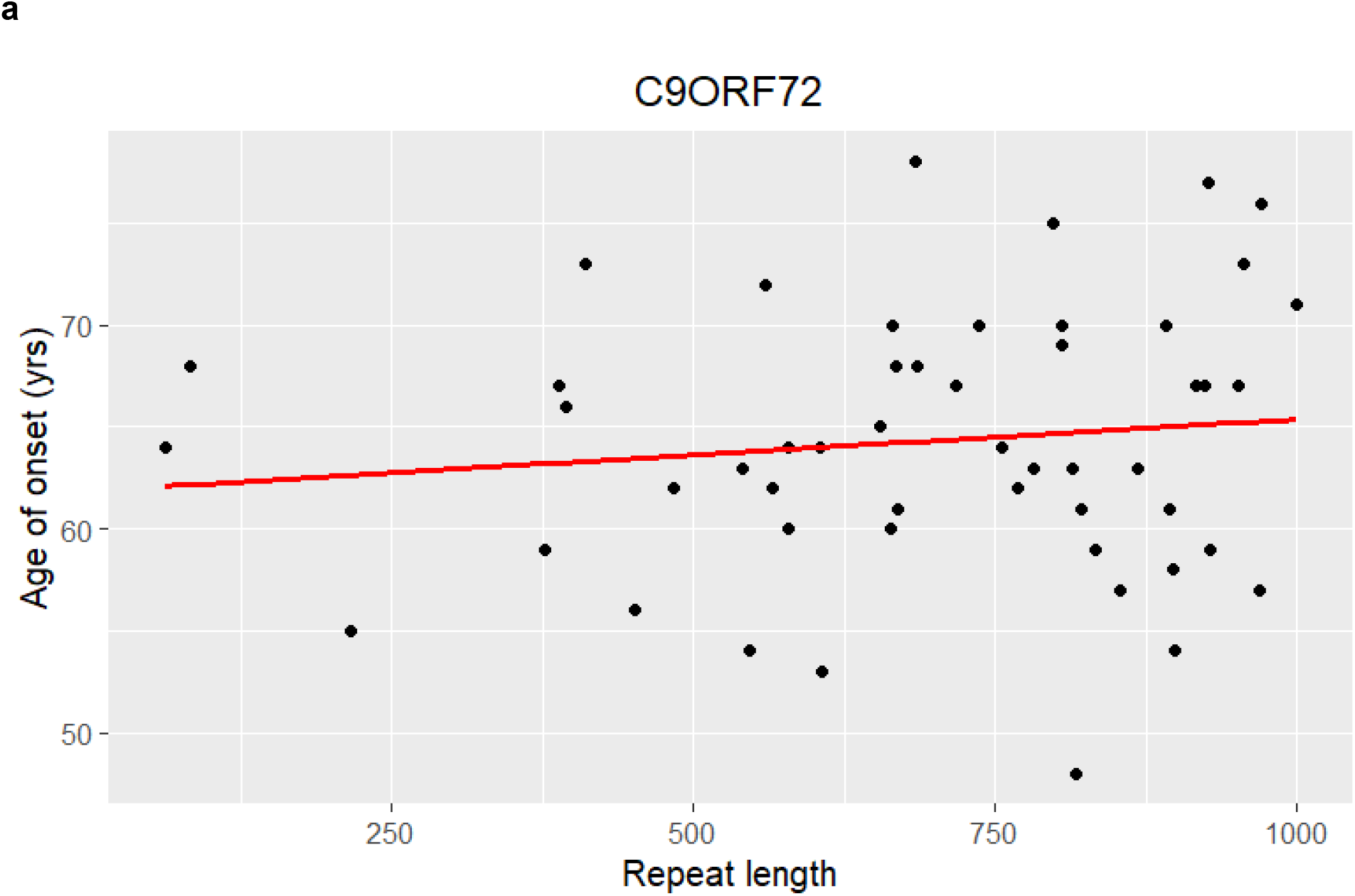
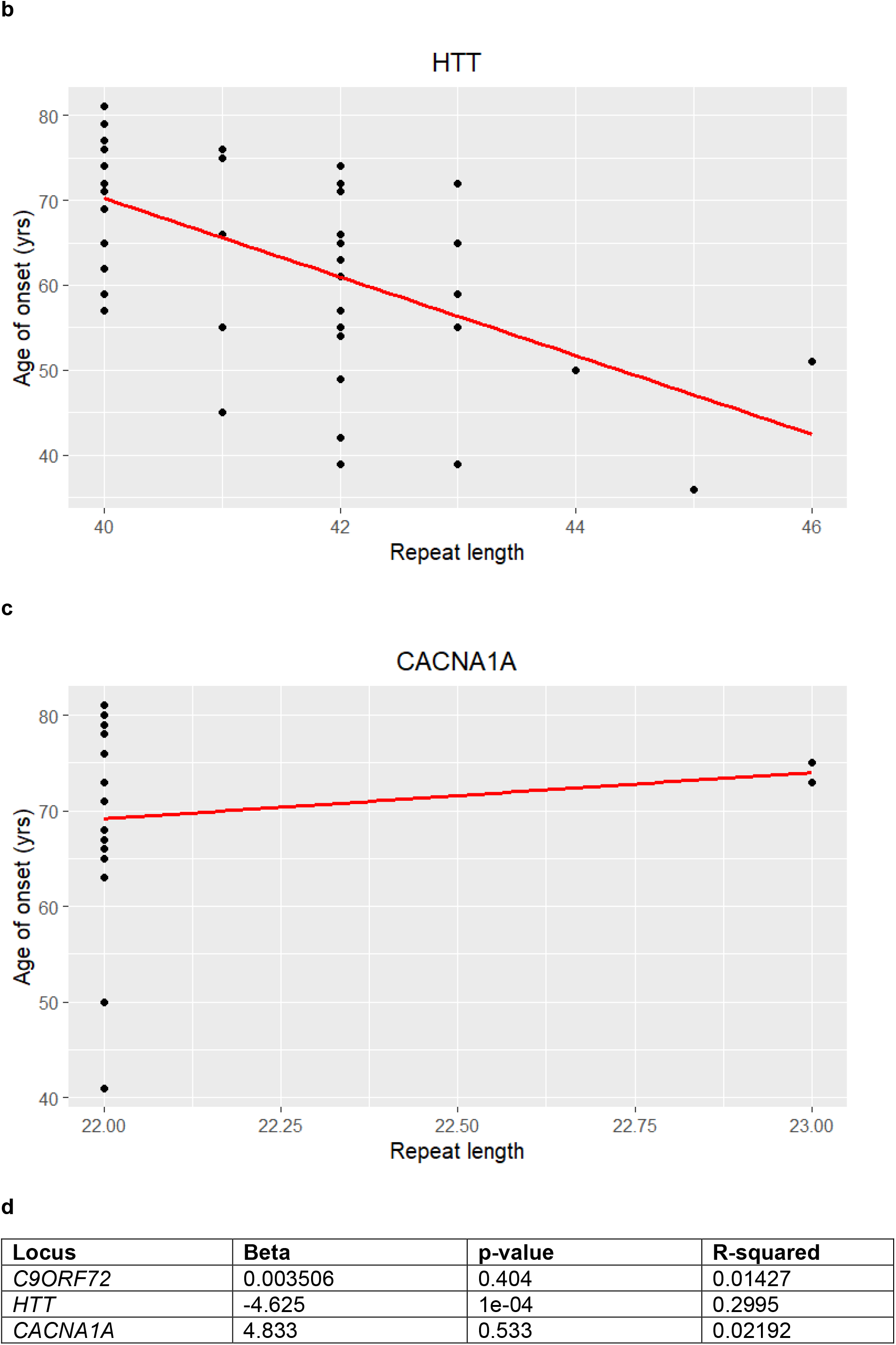
Age of onset by function of the repeat size. **(a)** Relationship between age when motor neuron disease was first reported and repeat size for the *C9ORF72* locus; **(b)** Relationship between age at HD first occurrence and repeat size for the *HTT* locus; **(c)** Relationship between age at SCA first occurrence and repeat size for the *CACNA1A* locus. In red, linear model for this relationship. **(d)** Coefficients of a linear model of the relationship between age of onset and repeat size for each locus.

**Supplementary Figure 4.**
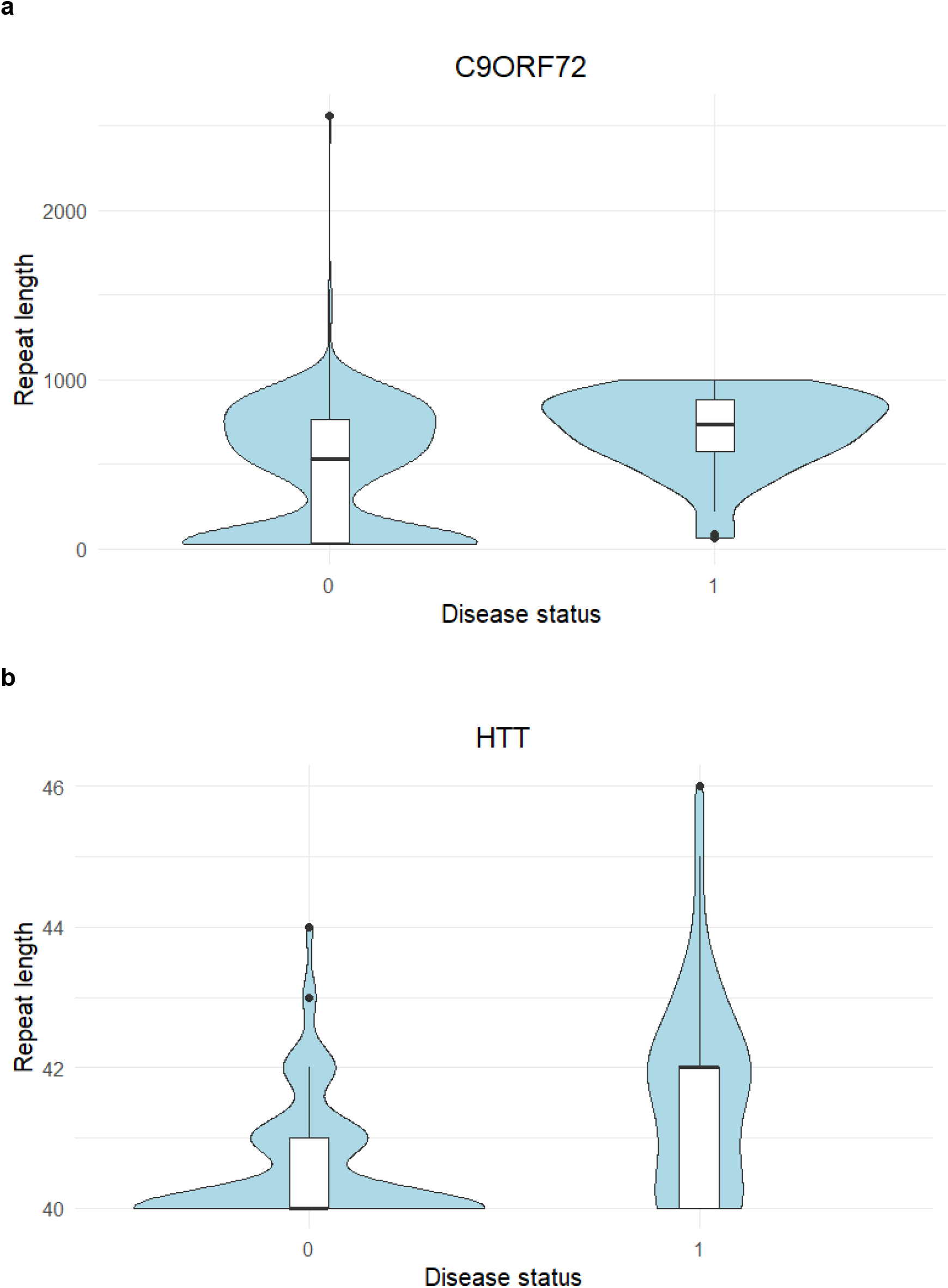
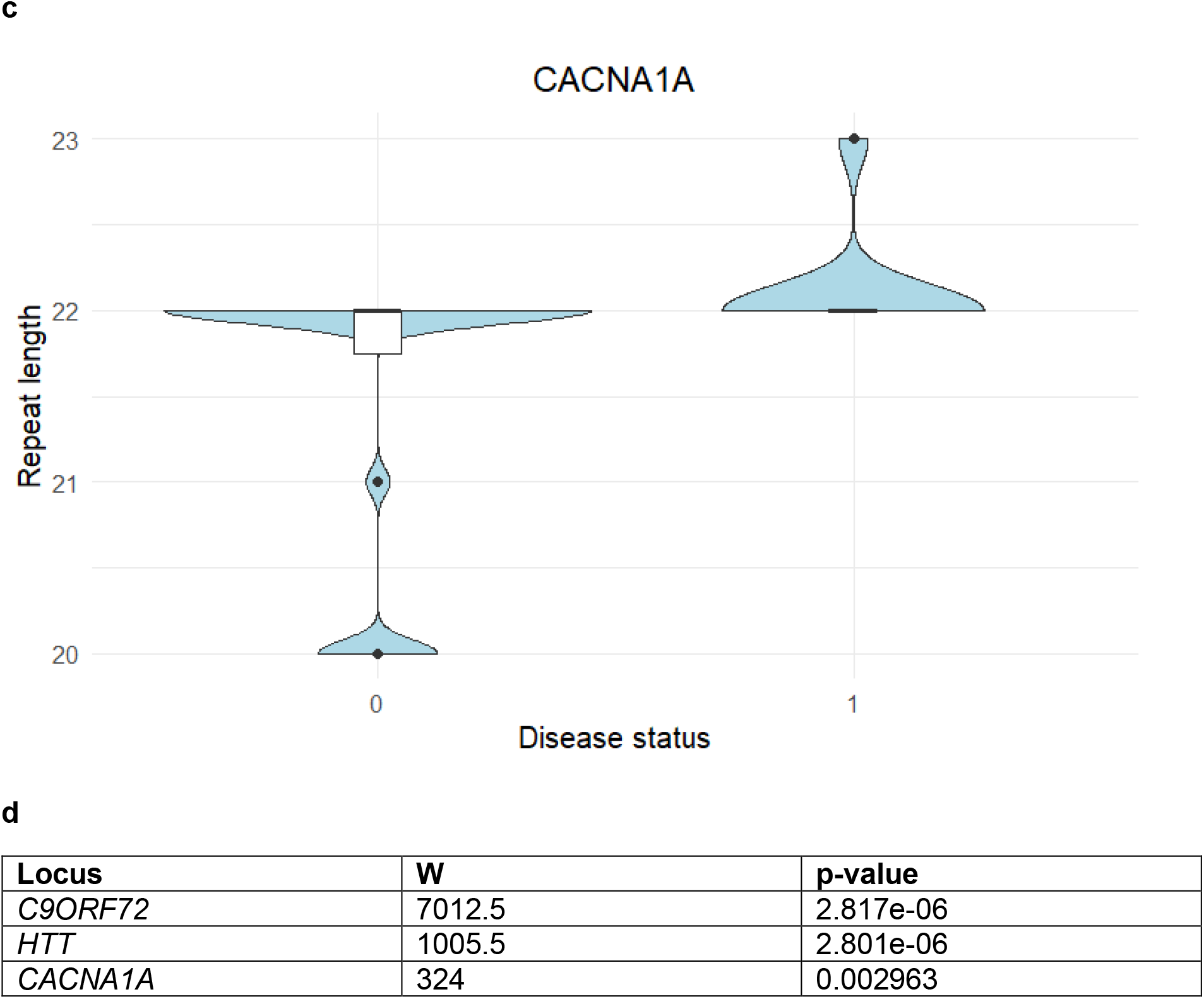
Distribution of repeat sizes for each locus by function of the disease status. **(a**) Distribution of *C9ORF72* repeat sizes among individuals diagnosed with motor neuron disease and individuals that remained symptom-free; **(b)** Distribution of *HTT* repeat sizes among individuals diagnosed with HD and individuals that remained symptom-free; **(c)** Distribution of *CACNA1A* repeat sizes among individuals diagnosed with SCA and individuals that remained symptom-free. Disease status 0 = Symptom-free, Disease status 1 = Diagnosed. **(d)** Summary of Wilcoxon rank sum test statistics.

**Supplementary Table 1.**
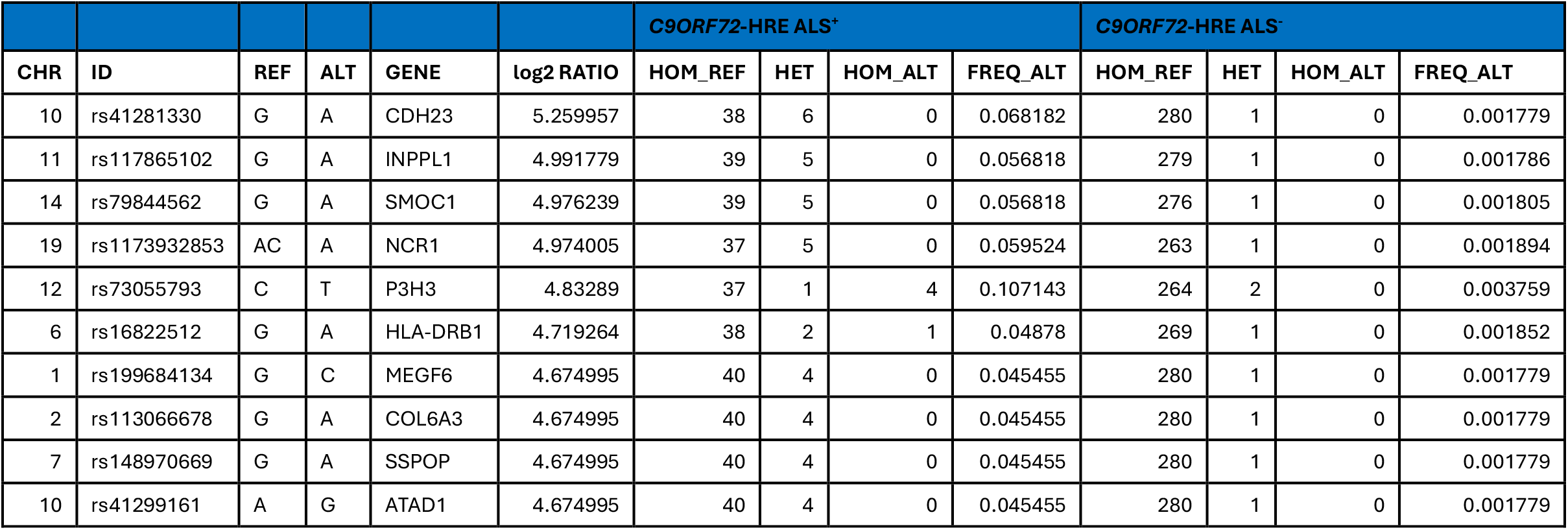
Top 10 enriched variants in the *C9ORF72*-HRE ALS^+^ group vs the *C9ORF72*-HRE ALS^−^ group. CHR, Chromosome; ID, Variant ID; REF, Reference allele; ALT, Alternative allele; GENE, Gene symbol; log2 RATIO, log2 of the ratio between the alternative allele frequency in the *C9ORF72*-HRE ALS^+^ group and the alternative allele frequency in the *C9ORF72*-HRE ALS^−^ group; HOM_REF_CTS, Reference allele homozygous counts; HET_REF_ALT_CTS, Heterozygous counts; TWO_ALT_GENO_CTS; Alternative allele homozygous counts; FREQ_ALT, Frequency of the alternative allele. (Autosomal variants only).

## Notes

### Funding Statement

This study was funded by Eli Lilly and Company.

### Author Declarations

The UK Biobank database received ethical approval from the North-West Haydock Research Ethics Committee (REC reference 21/NW/0157) and participants gave informed consent.

